# Improving SARS-CoV-2 cumulative incidence estimation through mixture modelling of antibody levels

**DOI:** 10.1101/2021.04.09.21254250

**Authors:** C. Bottomley, M. Otiende, S. Uyoga, K. Gallagher, E.W. Kagucia, A.O. Etyang, D. Mugo, J. Gitonga, H. Karanja, J. Nyagwange, I.M.O. Adetifa, A. Agweyu, D.J. Nokes, G.M. Warimwe, J.A.G. Scott

## Abstract

As countries decide on vaccination strategies and how to ease movement restrictions, estimates of cumulative incidence of SARS-CoV-2 infection are essential in quantifying the extent to which populations remain susceptible to COVID-19. Cumulative incidence is usually estimated from seroprevalence data, where seropositives are defined by an arbitrary threshold antibody level, and adjusted for sensitivity and specificity at that threshold. This does not account for antibody waning nor for lower antibody levels in asymptomatic or mildly symptomatic cases. Mixture modelling can estimate cumulative incidence from antibody-level distributions without requiring adjustment for sensitivity and specificity. To illustrate the bias in standard threshold-based seroprevalence estimates, we compared both approaches using data from several Kenyan serosurveys. Compared to the mixture model estimate, threshold analysis underestimated cumulative incidence by 31% (IQR: 11 to 41) on average. Until more discriminating assays are available, mixture modelling offers an approach to reduce bias in estimates of cumulative incidence.

**One-Sentence Summary:** Mixture models reduce biases inherent in the standard threshold-based analysis of SARS-CoV-2 serological data.

## Main Text

Together with the total number of cases and deaths, the cumulative incidence of infection is a key parameter for assessing the impact of the SARS-CoV-2. The evidence to date suggests that reinfection is uncommon, at least in the short term, and associated with mild disease (1,2). Therefore, estimating cumulative incidence—the proportion that have been infected—can help to establish to what extent populations are protected from severe disease by natural infection.

Given the substantial under-ascertainment through PCR testing, data on antibody levels provide the most reliable means of estimating cumulative incidence. The conventional analysis of these data involves estimating the proportion that lies above an arbitrary threshold and adjusting for the sensitivity and specificity at that threshold (3,4). However, sensitivity is usually estimated using samples from PCR-positive cases who are symptomatic and have been recently infected. Since these samples are likely to have higher antibody levels than samples from the general population of infected individuals, including those with asymptomatic infection, this can lead to spectrum bias and the overestimation of sensitivity (5). Mixture models offer an alternative for estimating cumulative incidence from serological data that does not involve specifying a threshold and is therefore not vulnerable to spectrum bias (6,7).

We used data from surveys of Kenyan blood donors (4,8), healthcare workers (9), truck drivers/assistants (10) and pregnant women (11) to compare the standard threshold-based approach to cumulative incidence estimation with the mixture modelling approach. Through this comparison we are able to illustrate the degree of bias in standard seroprevalence estimates.

Across all serosurveys, the samples were tested for anti-SARS-CoV-2 IgG antibodies using an adaptation of the Krammer ELISA for whole length spike antigen (12). Ratios of optical densities (OD) relative to a negative control were used to quantify the antibody concentrations. The assay was originally validated using 910 pre-COVID serum samples collected in 2018, all of which were collected from adults and children from the Coast region of the country, and samples from 174 PCR-positive Kenyan adults, which were collected from patients admitted to Kenyatta National Hospital in Nairobi and their contacts (14 pre-symptomatic, 55 symptomatic, 92 asymptomatic and 13 unknown). The validation was based on a threshold OD ratio of 2, and yielded sensitivity and specificity estimates of 92.7% and 99.0% respectively. The performance of the assay was found to be consistent with that of other assays in a WHO-sponsored international standardization study (13).

To estimate cumulative incidence by the threshold approach we used a Bayesian model to adjust for sensitivity and specificity at the selected threshold (14). For the mixture model approach we assumed a normal distribution of log-scale antibody concentrations in uninfected individuals and a skew normal distribution in infected individuals (7). In this model, the proportion infected—i.e. the cumulative incidence— is one of the parameters that defines the mixture model, and is therefore estimated as part of the model fitting process. Since we anticipated that the two distributions would be overlapping, and mixture models can be difficult to fit in these circumstances, we imposed several constraints as described in the Supplementary Material. In particular, we fixed the standard deviation (SD) of the uninfected (normal) distribution to be equal to the observed SD in the pre-COVID-19 samples. By fixing the SD in this way, we are effectively assuming that excess variation—i.e. variation beyond what is predicted for the uninfected population—is attributable to infection. Both models were fitted using the Rstan package in R (15,16).

The distributions estimated from the mixture model (Figure 2) did not segregate as clearly as expected based on the distributions for pre-COVID-19 and PCR-positive samples (Figure 1). This was mainly because the distribution of antibody-levels in the infected component was shifted to the left relative to the distribution in PCR positive samples, i.e. the means were significantly lower in magnitude than the mean in the PCR positive samples (mean log_2_ OD ratio = 3.07, Supplementary Table 2). In contrast, the mean in the uninfected component was usually close to the mean observed in the pre-COVID-19 samples (mean log_2_ OD ratio = -0.17), though the proximity of the uninfected mean to the pre-COVID-19 mean varied between surveys; for example, in truck drivers the means were higher than the pre-COVID-19 mean and in pregnant women they were lower. In most surveys, the skew parameter was close to zero and the scale parameter, which determines the spread of the distribution, was similar to the standard deviation in PCR positive cases (log_2_ scale SD = 1.32).

**Figure 1:**
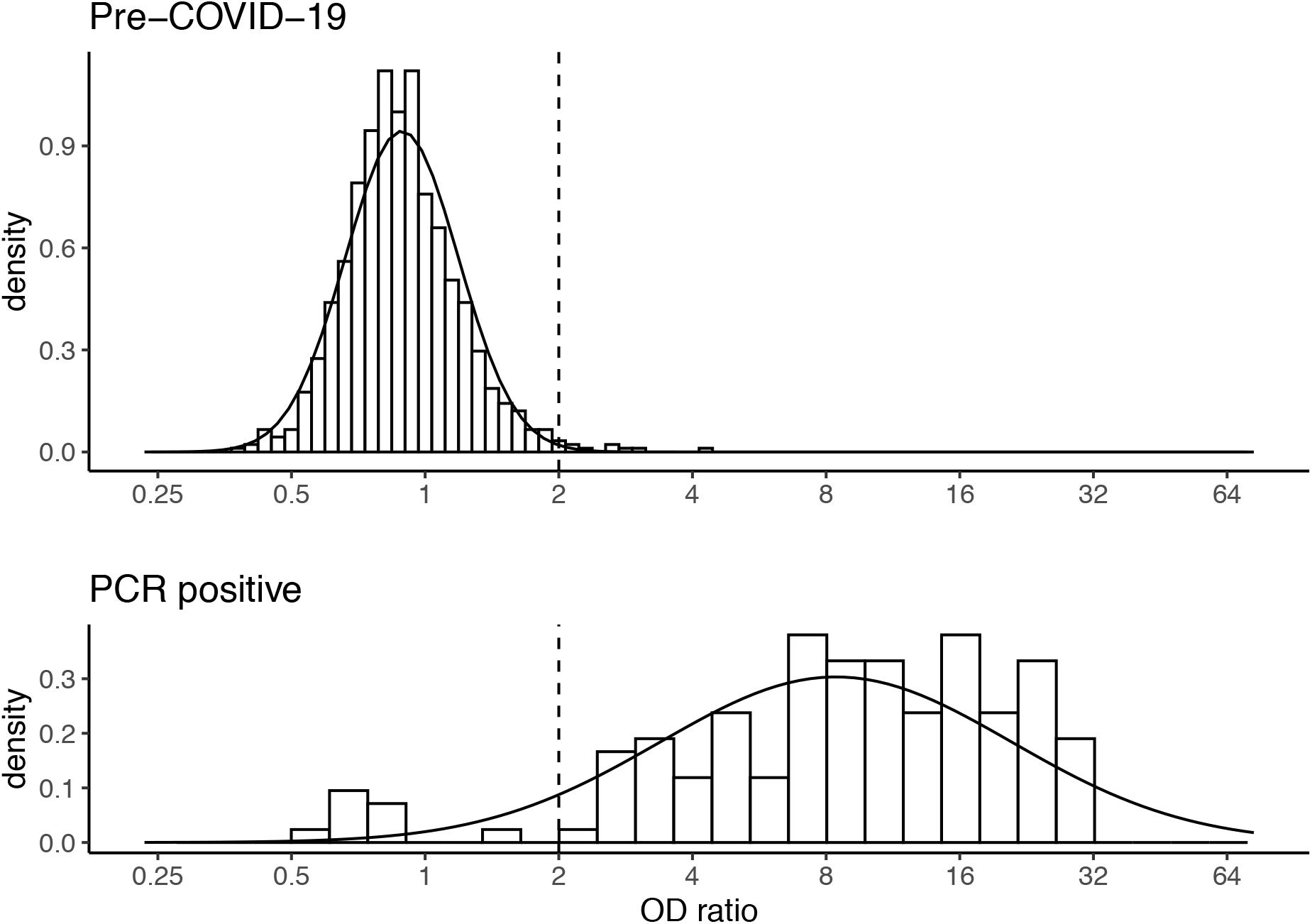
Distribution of anti-spike IgG antibodies in PCR positive samples and pre-COVID samples. The dotted line indicates the threshold (OD ratio > 2) used to define seropositivity.

**Figure 2:**
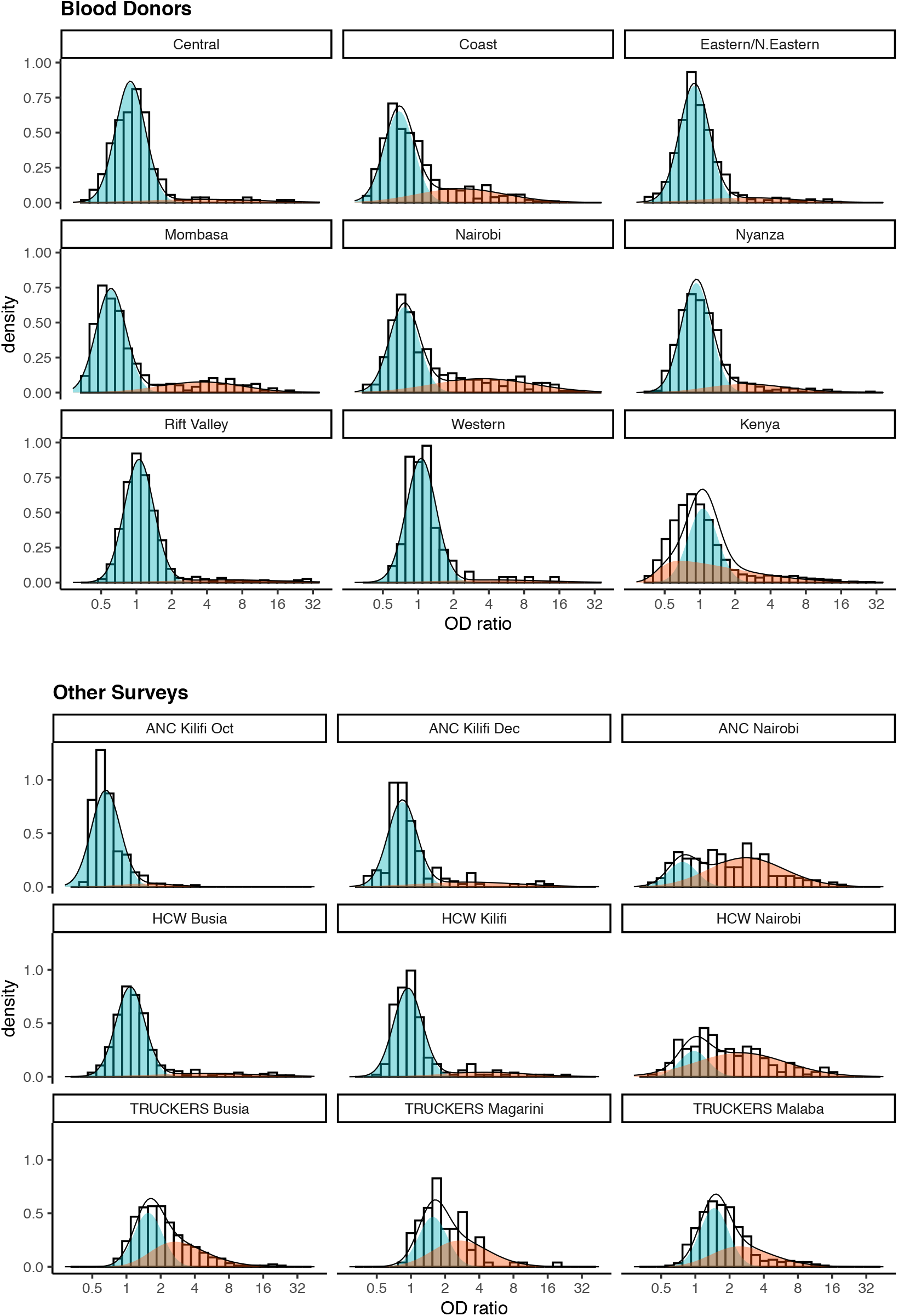
Mixture distributions fitted to anti-spike IgG antibody data collected in serological surveys of Kenyan blood donors, antenatal care (ANC) attendees, healthcare workers (HCW), and truck drivers. The red distributions represent responses in individuals who have been exposed to previous SARS-CoV-2 infection and the blue distributions represent responses in uninfected individuals.

Mixture model-derived estimates of cumulative incidence were higher than standard threshold-based estimates in almost all surveys. On average (median), the mixture model estimate was 1.44-fold higher than the threshold-based estimate (IQR: 1.12, 1.70). In other words, the threshold analysis underestimated cumulative incidence by 31% (IQR: 11 to 41), assuming that the mixture model estimates are unbiased. In the analysis of blood donor samples from the Coast region—the analysis that required least extrapolation because the SD for the uninfected component was estimated using pre-COVID-19 samples from this region—there was an almost 2-fold relative difference (31% versus 16%). In general, the associated 95% credible intervals tended to be wider for the mixture model estimate, with the largest difference occurring when there was strong overlap between the component distributions (Figure 3). For example, the three surveys of truck drivers (in Busia, Magarini and Malaba) produced the widest confidence intervals; they were also the surveys with the greatest overlap between the distributions.

**Figure 3:**
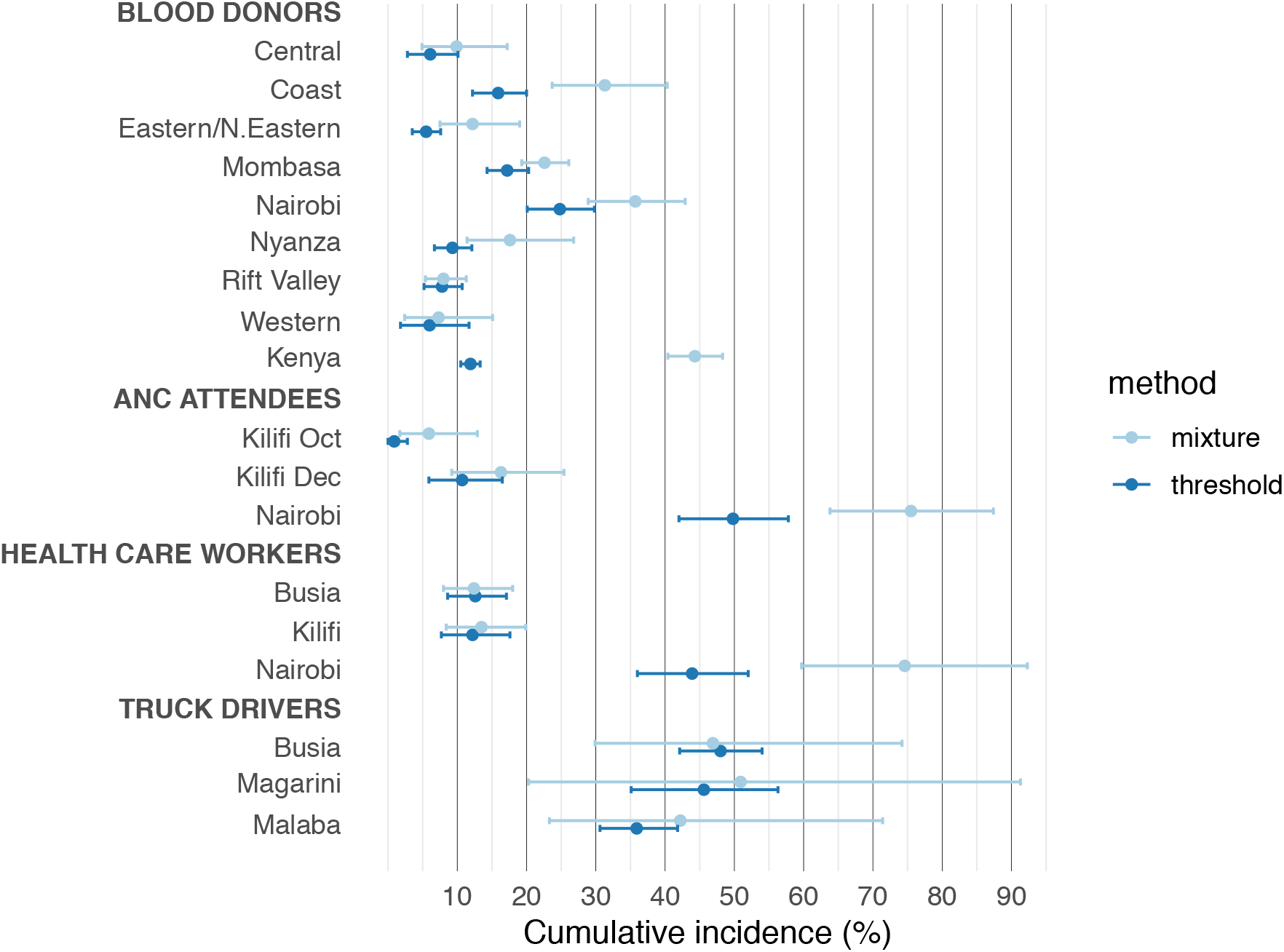
Comparison of SARS-CoV-2 cumulative incidence estimates obtained using the standard threshold-based approach (adjusted for sensitivity and specificity) with those obtained using the mixture model.

Overall, the mixture models fitted the data well (Figure 2) and produced plausible parameter estimates. A notable exception was the countrywide analysis of blood donor data. Here the fit was poor, and the resulting cumulative incidence estimate was implausibly high. Furthermore, the mean of the uninfected distribution was lower than in other analyses, and the skew and scale parameter estimates were higher.

To further explore the issue of misspecification and assess the robustness of the mixture model, we tested the model in three scenarios (Supplementary Table 3). In the first scenario we combined data from the PCR positives and pre-COVID samples and fitted the mixture model to these combined data. In the second scenario, we assumed the distribution in infected individuals is itself a mixture. The components of this mixture might correspond, for example, to first time infections and reinfections. Data were simulated for this scenario using a 3-component normal mixture model, with component 1 representing the distribution in uninfected individuals and components 2 and 3 representing the distribution in infected individuals. Finally, in the third scenario, we assumed a mixture distribution in the uninfected population. This scenario was designed to inform analyses where data are combined from different populations with varying background antibody levels, such as in the Kenya-wide analysis of blood donors. Data were again simulated from a mixture of three normal distributions, but this time with components 1 and 2 representing the distribution in uninfected individuals and component 3 representing the distribution in infected individuals.

The mixture model performed well in scenarios 1 and 2. In both cases estimates of the proportion infected were close to the expected values (scenario 1: 15%, vs 14%; scenario 2: 19% vs 20%). However, in scenario 3 the mixture model estimate was severely biased (estimated = 36%, expected = 20%). The poor performance of the mixture model in this scenario is likely to be due to the assumed SD. The SD should reflect total variation in the uninfected subpopulation. However, in scenario 3 it does not account for the between-subgroup variation in the uninfected population. Consequently, the mixture model is forced to attribute the excess variation to the infected component thereby overestimating the proportion infected. This scenario arises in the Kenya-wide analysis; the pre-COVID-19 samples were collected from a single region and therefore the assumed SD of the uninfected does not account sufficiently for the between-region variation in background antibody levels.

In general, the mixture modelling suggests that, on average, antibody levels in infected serosurvey participants are significantly lower than in the PCR-positive samples used to estimate sensitivity. Because of this, threshold-based estimates— which rely on accurate sensitivity estimates—will underestimate cumulative incidence.

There are two potential explanations for the lower antibody levels in these populations compared to PCR positives. First, serosurvey participants are more likely to be asymptomatic or mildly symptomatic cases who have lower antibody levels than symptomatic cases (17–19). Cough, fever and hospitalisation are all positively associated with higher antibody levels (20). Second, the samples used to estimate sensitivity are generally obtained from recently infected individuals who are likely to have high antibody levels. In the PCR-positive samples used in our study, the median time between symptom onset and blood sample collection was 21 days, which may be ideal to capture the peak antibody response.

Antibody waning has been reported in a number of studies (17–19,21,22). Although waning is generally greater for anti-nucleocapsid protein antibodies it is also significant for anti-spike protein antibodies, particularly among milder cases of infection where the antibody half-life has been estimated to be 73 days (23). Of the two explanations, waning appears to be the more important in our study because there was only a modest difference in antibody levels between asymptomatic and symptomatic cases (mean log_2_ OD ratio: 2.9 vs 3.4). Moreover, studies that have specifically accounted for waning by assuming a constant rate of seroreversion, rather than by accounting for spectrum bias more generally as we have done, have also predicted that waning has a significant impact on estimates of cumulative incidence (19,24).

The mixture model results suggest variability in the background levels of anti-spike IgG between different populations. In addition to significant variation by region, we observed higher IgG levels in truck drivers and lower levels in pregnant women. Both in our analysis of Kenyan serological data and in our simulation example, we have shown that not accounting for this variation can produce biased estimates if data from different populations are combined. This variability also has the potential to introduce bias in seroprevalence estimates using a threshold-based analysis. For example, in a population with low background IgG levels, as observed in pregnant women, we anticipate that the specificity estimate will be too low and the sensitivity estimate too high.

The reasons for the variation in baseline IgG levels are unclear. It could simply reflect day-to-day or longitudinal temporal variation in the laboratory procedures, though the negative control should guard against this bias. Alternatively, it could be related to differences between populations in exposure to infection and possibly also infective dose. Several studies have reported variation in antibody levels between populations in samples collected pre-COVID-19, with antibody levels generally being higher in African populations than non-African populations (25,26). Furthermore, anti-SARS-CoV-2 antibodies are known to cross-react with antibodies against other coronaviruses (27), and possibly also antibodies against dengue (28) and malaria (29), though the latter finding was not confirmed in a more recent study (30). For pregnant women, it is possible that low antibody levels are a feature of immune environment in pregnancy (31).

The major limitation of the mixture model approach to cumulative incidence estimation is that it is sensitive to the variance assumed for uninfected population. Ideally the variance should be estimated using pre-COVID-19 samples from the population being surveyed, but in practice the two populations are often unrelated. For example, we used samples from pre-COVID-19 in the Coast region to estimate the variance, but then used this estimate to infer cumulative incidence in other populations, including blood donors in other regions and in different subgroups such as pregnant women. Another limitation of the mixture modelling approach is that it is necessary to assume parametric models for the component distributions. In our analysis we assumed the distribution among uninfected individuals was normal and the distribution among infected individuals was skew normal. However, we believe these distributional assumptions are probably of secondary concern. The data from pre-COVID-19 samples suggests that antibody levels in uninfected individuals are approximately normally distributed. And although we were unable to determine the distribution in infected individuals, our analysis of simulated data suggests that estimates of cumulative incidence are robust to misspecification of this distribution. Finally, we note that our analysis was limited to Kenyan serosurveys; in the future it will be important to explore using mixture models to analyse surveys that have been done elsewhere.

We have shown that cumulative incidence estimates based on a threshold analysis are likely to be biased downwards. While overestimating cumulative incidence can lead to complacency in the assessment of future COVID waves, underestimating cumulative incidence can also have serious adverse consequences if it prolongs social restrictions unnecessarily. This is particularly relevant in LMICs where resumption of educational, social and economic activities is unlikely to be brought about by rapid dissemination of COVID-19 vaccines. Here we provide an alternative to the standard threshold analysis for estimating cumulative incidence that does not require specific adjustments for waning and allows for differences between populations in background antibody levels. The approach makes assumptions about variation in background antibody levels that need to be validated locally, but until we have a better understanding of the spectrum of antibody concentrations by symptom severity, or due to waning, it is probably the more accurate of the two approaches for estimating cumulative incidence.

## Data Availability

The data are available on request from the corresponding author

## Acknowledgements

We thank the serosurvey participants for their contribution to this research.

## Funding

UK Medical Research Council (MRC) and the UK Department for International Development (DFID) under the MRC/DFID Concordat agreement, which is also part of the EDCTP2 programme supported by the European Union; grant reference: MR/R010161/1 (CB).

## Author contributions

Conceptualization: AS, CB

Methodology: CB, MO, AS

Investigation: SU, KG, EK, AE, DM, JG, HK, JN, IA, AA, GW

Writing – original draft: CB

Writing – review & editing: CB, MO, KG, EK, AE, IA, AA, DN, AS

## Competing interests

Authors declare that they have no competing interests

## Data and materials availability

All data and code are available in the supplementary materials.

## Supplementary Materials

Supplementary Text

Tables S1 to S3

Stan Code

## Notes

### Competing Interest Statement

The authors have declared no competing interest.

### Author Declarations

The component studies were approved by Kenya Medical Research Institute Scientific and Ethics Review Unit (Protocol SSC 3426 for blood donors and Protocol SSC 4085 for health care workers, antenatal care attendees and truck drivers). Further ethical approval for the study of antenatal care attendees was obtained from the Kenyatta National Hospital University of Nairobi Ethics Review Committee (Protocol P327/06/2020) and the Kilifi County health management rapid response team.

