## supplementary text and tables for "Improving SARS-CoV-2 cumulative incidence estimation through mixture modelling of antibody levels"

This file includes:

Supplementary Text

Tables S1 to S3

Stan Code

**Supplementary Text:**

*Sensitivity and specificity adjusted threshold analysis*

We incorporated information on the sensitivity and specificity of the threshold by simultaneously modelling the serosurvey data and validation data. Specifically, we modelled counts of i) the number, $y,$ of survey samples above the cut-off OD ratio, ii) the number, $x$, of PCR positive samples above the cut-off and iii) the number, $z,$ of pre-COVID-19 samples below the cut-off. We assumed a model where each count follows a binomial distribution

$y\sim\text{Binomial}\left( p_{obs},N \right).$

$$x\sim\text{Binomial}(se,N_{se})$$

$$z\sim\text{Binomial}\left( sp,N_{sp} \right)$$

and where the proportion seropositive (OR ratio > 2) is related to the cumulative incidence, $p,$and sensitivity, $se,$ and specificity, $sp$, according to the equation:

$$p_{obs}=se\times p+\left( 1-sp \right)\times\left( 1-p \right).$$

To fit the model, we used a uniform(0,1) prior for each parameter ($p,se,sp$).

*Description of the mixture model and priors*

We fitted a two-component mixture model where individuals are classified as either uninfected or as having experienced SARS-CoV-2 infection. We assumed that log${}_{2}$ OD ratios follow a skew normal among infected individuals (parameters: location = $\xi$, scale = $\omega$ and skew=$\alpha$) and a normal distribution among uninfected individuals. According to this model, the density across the population as a whole is

$$p\times\text{Skew Normal}(\xi,\omega,\alpha)+(1-p)\times\text{Normal}(\theta,\nu),$$

where $p$ is the cumulative incidence of SARS-CoV-2.

To make it easier to interpret the model parameters, we reparameterised in terms of the difference, $\delta$ , in the means of the two distributions:

$$\xi=\theta+\delta-\omega\sqrt{2/\pi}\frac{\alpha}{\sqrt{(1+\alpha^{2})}}$$

Several constraints were used to facilitate model fitting. First, we fixed the model standard deviation in the uninfected, $\nu,$ to be equal to the standard deviation in the pre-COVID-19 samples. Second, we used an informative prior for $\delta$ to ensure that $\delta>0$ and also that the magnitude of the difference was unlikely to exceed the difference between symptomatic PCR positive samples and pre-COVID-19 samples. Finally, we also used an informative prior to rule out strong skew in either direction.

Model priors:

$$\theta\sim\text{Normal}(0,10)$$

$$\delta\sim\text{Normal}^{+}(0,1.83)$$

$$\text{log }\omega\sim\text{Normal}(0,10)$$

$$\alpha\sim\text{Normal}(0,1)$$

$$p\sim\text{Uniform}(0,1)$$

Non-informative priors were used for $p$, $\theta$ and $\omega$ and informative priors were used for $\alpha$ and $\delta$. The prior for $\delta$ puts 5% probability on the difference in means exceeding the difference between symptomatic PCR positive cases and pre-COVID sample (log${}_{2}$ OD ratio = 3.60).

*Ethical Considerations*

The component studies were approved by Kenya Medical Research Institute Scientific and Ethics Review Unit (Protocol SSC 3426 for blood donors and Protocol SSC 4085 for health care workers, ANC attendees and truck drivers). Blood donors gave individual written consent for the use of their samples for research. Health care workers and truck drivers provided written and/or verbal informed consent for participation in the study. ANC attendees provided consent to have blood samples taken but did not explicitly consent to participate in research. Further ethical approval for the use of these samples was therefore sought and obtained from the Kenyatta National Hospital – University of Nairobi Ethics Review Committee (Protocol P327/06/2020) and the Kilifi County health management rapid response team.

**Supplementary Table S1:** Estimates of cumulative incidence

|  | **Survey period** | **N** | **OD ratio**  **> 2** | **Sensitivity & specificity adjusted** | | **Mixture model** | |
| --- | --- | --- | --- | --- | --- | --- | --- |
| **Kenyan blood donors** | 20 Aug - 30 Sep |  | % | % | 95% CI | % | 95% CI |
| Central |  | 225 | 6.2 | 6.1 | 2.8 - 10.1 | 9.9 | 4.9 - 17.2 |
| Coast |  | 435 | 15.4 | 15.9 | 12.2 - 20.0 | 31.3 | 23.7 - 40.3 |
| Eastern/ N. Eastern |  | 702 | 6.0 | 5.5 | 3.5 - 7.6 | 12.2 | 7.5 - 19 |
| Mombasa |  | 802 | 16.7 | 17.2 | 14.3 - 20.3 | 22.6 | 19.3 - 26.1 |
| Nairobi |  | 361 | 23.5 | 24.8 | 20.1 - 29.8 | 35.7 | 28.9 - 42.9 |
| Nyanza |  | 584 | 9.4 | 9.3 | 6.7 - 12.1 | 17.6 | 11.4 - 26.8 |
| Rift Valley |  | 508 | 8.1 | 7.8 | 5.2 - 10.7 | 8.0 | 5.4 - 11.3 |
| Western |  | 106 | 5.7 | 6.0 | 1.8 - 11.7 | 7.3 | 2.4 - 15.1 |
| All Regions |  | 3,723 | 11.9 | 11.9 | 10.5 - 13.3 | 44.3 | 40.4 - 48.3 |
| **Antenatal care attendees** |  |  |  |  |  |  |  |
| Kilifi | 18 Sep – 23 Oct | 264 | 1.1 | 0.9 | 0 - 2.8 | 5.9 | 1.7 - 12.9 |
| Kilifi | 24 Oct – 24 Nov | 155 | 10.3 | 10.7 | 5.9 - 16.5 | 16.3 | 9.2 - 25.4 |
| Nairobi | 30 Jul – 25 Aug | 196 | 46.4 | 49.8 | 42.0 - 57.8 | 75.5 | 63.8 - 87.4 |
| **Healthcare workers** |  |  |  |  |  |  |  |
| Busia | 19 Oct – 23 Oct | 301 | 12.3 | 12.6 | 8.6 - 17.1 | 12.4 | 8.0 - 18.0 |
| Kilifi | 13 Oct – 4 Dec | 200 | 11.5 | 12.2 | 7.7 - 17.6 | 13.5 | 8.4 - 19.9 |
| Nairobi | 30 July – 25 Aug | 183 | 41.0 | 43.9 | 36.0 - 52.0 | 74.6 | 59.7 - 92.3 |
| **Truck drivers** |  |  |  |  |  |  |  |
| Busia | 13 – 15 Oct | 365 | 44.7 | 48.0 | 42.1 - 54.0 | 46.9 | 29.9 - 74.2 |
| Magarini | 30 Sep – 23 Oct | 101 | 42.6 | 45.6 | 35.1 - 56.3 | 50.9 | 20.3 - 91.3 |
| Malaba | 13 – 15 Oct | 364 | 33.8 | 35.9 | 30.6 - 41.8 | 42.2 | 23.3 - 71.4 |

**Supplementary Table S2:** Mixture model parameter estimates

|  | **SARS-CoV-2 Uninfected:** | | **SARS-CoV-2 Infected:** | | | | | |
| --- | --- | --- | --- | --- | --- | --- | --- | --- |
|  | **Mean** | | **Mean** | | **Scale** | | **Skew** | |
| **Kenyan blood donors** | estimate | 95% CI | estimate | 95% CI | estimate | 95% CI | estimate | 95% CI |
| Central | -0.17 | -0.24 , -0.11 | 1.76 | 0.76 , 2.79 | 1.70 | 1 , 2.67 | 0.11 | -1.79 , 2.13 |
| Coast | -0.54 | -0.60 , -0.48 | 1.05 | 0.70 , 1.43 | 1.34 | 1.02 , 1.79 | 0.45 | -1.15 , 2.46 |
| Eastern/ N. Eastern | -0.16 | -0.20 , -0.12 | 1.16 | 0.64 , 1.77 | 1.49 | 1.09 , 2.01 | 0.49 | -1.35 , 2.59 |
| Mombasa | -0.72 | -0.76 , -0.69 | 1.95 | 1.70 , 2.18 | 1.43 | 1.04 , 1.93 | -0.96 | -2.61 , 0.79 |
| Nairobi | -0.39 | -0.46 , -0.33 | 1.71 | 1.37 , 2.05 | 1.47 | 1.16 , 1.95 | 0.30 | -1.23 , 2.13 |
| Nyanza | -0.11 | -0.15 , -0.06 | 1.21 | 0.74 , 1.71 | 1.50 | 1.1 , 1.93 | 1.24 | -0.86 , 3.11 |
| Rift Valley | 0.07 | 0.03 , 0.11 | 2.33 | 1.68 , 2.97 | 1.68 | 1.18 , 2.38 | 0.27 | -1.49 , 2.26 |
| Western | 0.08 | -0.01 , 0.17 | 2.07 | 0.65 , 3.35 | 1.56 | 0.57 , 3.31 | 0.01 | -1.89 , 2.01 |
| All Regions | -0.25 | -0.28 , -0.22 | 0.45 | 0.37 , 0.54 | 2.12 | 2.02 , 2.23 | 6.76 | 5.61 , 7.93 |
| **Antenatal care attendees** | |  |  |  |  |  |  |  |
| Kilifi, Oct | -0.61 | -0.67 , -0.55 | 0.55 | -0.12 , 1.33 | 0.82 | 0.44 , 1.41 | 0.02 | -0.96 , 1.00 |
| Kilifi, Dec | -0.25 | -0.33 , -0.17 | 1.48 | 0.76 , 2.24 | 1.55 | 1.02 , 2.3 | 0.36 | -1.53 , 2.36 |
| Nairobi | -0.38 | -0.58 , -0.19 | 1.37 | 1.09 , 1.64 | 1.26 | 0.98 , 1.63 | 0.72 | -0.91 , 2.35 |
| **Healthcare workers** | |  |  |  |  |  |  |  |
| Busia | 0.11 | 0.06 , 0.16 | 2.22 | 1.53 , 2.88 | 1.62 | 1.12 , 2.35 | 0.18 | -1.65 , 2.19 |
| Kilifi | -0.09 | -0.15 , -0.02 | 2.05 | 1.39 , 2.60 | 1.31 | 0.85 , 1.99 | 0.25 | -1.54 , 2.21 |
| Nairobi | -0.04 | -0.33 , 0.24 | 1.17 | 0.86 , 1.49 | 1.43 | 1.13 , 1.87 | 0.50 | -1.09 , 2.37 |
| **Truck drivers** | |  |  |  |  |  |  |  |
| Busia | 0.64 | 0.51 , 0.75 | 1.57 | 1.19 , 1.94 | 1.17 | 0.89 , 1.42 | 1.88 | -0.11 , 3.42 |
| Magarini | 0.64 | 0.35 , 0.88 | 1.35 | 0.95 , 1.90 | 1.04 | 0.74 , 1.46 | 1.11 | -0.93 , 2.85 |
| Malaba | 0.55 | 0.42 , 0.69 | 1.28 | 0.90 , 1.74 | 1.20 | 0.88 , 1.53 | 1.48 | -0.78 , 3.41 |

**Supplementary Table S3:** Three tests of the mixture model

|  | **Data** | | | **Mixture model estimate** | | |
| --- | --- | --- | --- | --- | --- | --- |
|  | *Uninfected* | *Infected* | *% Infected* | *Uninfected* | *Infected* | *% Infected* |
| **Test 1**: PCR +ve (symptomatic and asymptomatic) and pre-COVID-19 samples combined | N = 910  Mean = -0.17  SD = 0.42 | N = 147  Mean = 3.07  SD = 1.32 | **14%** | Mean = -0.19 | Mean = 3.06  Scale = 1.72  Skew = -2.25 | **15%** |
| **Test 2**: Mixture of distributions in infected individuals (simulated data) | N = 200  Mean = 0  SD = 0.42 | N = 25, 25  Mean = 2, 0.5  SD = 1.4, 1.4 | **20%** | Mean = -0.01 | Mean = 1.37  Scale = 2.06  Skew = -0.29 | **19%** |
| **Test 3**: Mixture of distributions in uninfected individuals (simulated data) | N = 100, 100  Mean = -0.3, 0.3  SD = 0.42, 0.42 | N = 50  Mean = 1.4  SD = 1.4 | **20%** | Mean = -0.03 | Mean = 0.86  Scale = 1.67  Skew = 0.52 | **36%** |

**Stan Code: Threshold analysis adjusted for sensitivity and specificity**

data {

int N;

int N_se;

int N_sp;

int y;

int x;

int z;

}

parameters {

real<lower=0,upper=1> p;

real<lower=0,upper=1> se;

real<lower=0,upper=1> sp;

}

transformed parameters {

real<lower=0,upper=1> p_obs;

p_obs = se * p + (1 - sp) * (1 - p);

}

model {

//priors

p ~ beta(1, 1);

se ~ beta(1, 1);

sp ~ beta(1, 1);

//likelihood

y ~ binomial(N, p_obs);

x ~ binomial(N_se, se);

z ~ binomial(N_sp, sp);

}

**Stan Code: Mixture model**

data {

int N;

vector[N] y;

real nu;

}

parameters {

real theta;

real<lower=0> delta;

real log_omega;

real alpha;

real<lower=0,upper=1> p;

}

transformed parameters {

real xi;

real<lower=0> omega;

real theta_delta;

omega = exp(log_omega);

theta_delta = theta + delta;

xi = theta + delta - sqrt(2/3.142) * omega * alpha/sqrt(1 + alpha^2);

}

model {

// priors

theta ~ normal(0, 10);

delta ~ normal(0, 1.83);

log_omega ~ normal(0, 10);

p ~ beta(1,1);

alpha ~ normal(0, 1);

//likelihood

for (n in 1:N) {

target += log_sum_exp(log(p)

+ skew_normal_lpdf(y[n] | xi, omega, alpha),

log1m(p)

+ normal_lpdf(y[n] | theta, nu));

}

}
